# OPTIONAL VACCINE UTILIZATION HAS A MALE GENDER PREFERENCE – A RECORD BASED ANALYSIS FROM A TERTIARY CARE CENTRE OF EASTERN INDIA

**DOI:** 10.1101/2020.05.17.20072173

**Authors:** Sandeep Kumar Panigrahi, Anumita Maiti, Venkatarao Epari

## Abstract

**Background And Objectives:** Optional vaccines find an important place in immunization today. This study attempts to find out the trend of optional vaccine utilization, over the past three years in the immunization clinic of a tertiary care centre and to find out the association of gender disparity with the utilization of these vaccines.

**Methods:** The retrospective study was conducted during October to December 2016 using the register based secondary data of October 2013 to September 2016. Month wise utilization of optional vaccines *(Pneumococcal, influenza, typhoid, varicella, hepatitis A* and *MMR)* and *measles* was captured. Analysis was done using Stata 12.1 SE.

**Results:** An increasing trend of utilization was seen for all vaccines including optional vaccines. The mean doses received by male children was significantly more for all optional vaccines (unlike all vaccines taken together) as well as for individual vaccines like *Pneumococcal, influenza, typhoid, MMR, hepatitis A* (p<0.05), but not for *varicella* and *measles* vaccine (p>0.05).

**Conclusion:** Gender disparity (preference for male children) was present for all optional vaccines except *varicella*, and not in case of *measles* vaccine used in universal immunization program selected as control.

**Author’s note:**

What is already known on this subject
- Differences in uptake are related to various factors in national immunization program of India.
- Optional vaccines are not a part of the national immunization program,
- There is no clarity regarding pattern and trend of use and gender differences, if any for optional vaccines.

What this study adds
- Trend of optional vaccine uptake is on the rise reflecting increasing demand among parents.
- Gender differences do not exist for vaccines covered under national immunization program, as seen for *Measles* vaccine, probably because of universalization.
- Preference for male children exists for most of the optional vaccines (even for *MMR)* probably because of the cost involved.
- Gender disparity was absent in *Varicella*, probably because of fear of parents for scar marks on face of their girl child.

## INTRODUCTION

Immunization is one of the most effective public health strategies for restricting spread of various communicable diseases.(Patnaik, Mishra, and Choudhury 2014) In India, a child dies every minute from vaccine-preventable diseases. It also accounts for almost a fifth of all global under-five deaths; a large number of which are caused by vaccine-preventable diseases such as tuberculosis, pneumonia and diarrhoea. (United Nations Children’s Fund (UNICEF) 2016) This is a basic and fundamental preventive service provided free of charge at all public health care facilities in India. The benefits of immunization are not only restricted to improvement in health and life expectancy, but also have social and economic impact at both community and national level. Countries all over the world have their own national immunization program and agenda with an aim to cover some of the common life threatening diseases that can be prevented through these vaccines. Some countries have moved beyond the routine vaccines to vaccines meant to prevent meningococcal meningitis, cervical cancer, etc.

Universal Immunization program (UIP) in India, commonly known as Routine Immunization Program (RIP), aims to universally every mother and child. It currently comprises of the following vaccines: *Bacillus Calmette–Guérin (BCG)* for prevention of tuberculosis, *Oral Polio Vaccine (OPV)* and *Inactivated Polio Vaccine (IPV)* for poliomyelitis, *Pentavalent vaccine* (consisting of vaccines for prevention of Diphtheria, Pertussis, Tetanus, Hepatitis B and Haemophilus influenzae type b), *Measles Rubella (MR)* vaccine for Measles and Rubella prevention, *Diphtheria Pertussis and Tetanus (DPT)* vaccine used as booster, and Tetanus toxoid vaccine used as booster. *Rotavirus vaccine* for rotavirus associated diarrhoea, *Japanese encephalitis (JE) vaccine* for Japanese encephalitis and *Pneumococcal* vaccine for the pneumococcal pneumonia are given in selected states of the country. (Panigrahi and Mahapatro 2015; National Health Portal India 2018)

Vaccination coverage varies considerably from state to state. Differences in uptake are related to various factors like geographical distribution, regional trend, differences between rural and urban, poor and rich, and gender related factors. Complete immunization coverage in urban areas of Odisha was 68 percent and similar to rural Odisha.(Vital Statistics Division and Government of India 2014) Similar were the results when immunization coverage was assessed in an urban city of Odisha (Cuttack had 65 percent coverage) in 2012.(Prusty et al. 2013) This study is an attempt to find out the current trend of optional vaccines use over the past three years in the immunization clinic of a tertiary care centre in India and to find if there is any association of this trend for each vaccine with gender of the beneficiary.

## OBJECTIVES

1. To find out the trend of optional vaccine utilization, that are not covered in the national immunization schedule, over the past three years in the immunization clinic of a tertiary care centre.
2. To find out the association of gender with the observed trend of vaccination.

## METHODOLOGY

The study was a retrospective observational study based on secondary data available from the outpatient register of immunization and child health guidance clinic of the tertiary care centre in eastern part of India. The tertiary care centre is a teaching institute with outpatient turnover of as much as 1000-1200 patients each day. The data collection from registers and records available in the immunization and health guidance clinic was conducted from October to December 2016. The register and records from October 2013 to September 2016 were reviewed and data were collected. The sampling unit was a vaccine dose. Universal sampling method was used, and type of vaccine and gender related data for each vaccine was collected. Name of the vaccine and its type (e.g. PCV-13 is a pneumococcal vaccine, here PCV-13 is the name of the vaccine and pneumococcal vaccine is the type) of individual vaccine and gender of the recipient were considered for each dose. Month wise utilization of doses of vaccines (all vaccines, few optional vaccines and a control vaccine- *Measles*, given to everyone by government) were included in the data collection sheet. Tally marks were used to compile the data for each of the component variables needed for the study. Data were finally entered into an excel sheet.

Our study included those vaccines where a trend analysis was operationally feasible, and vaccines not facing stock out during this period. Optional vaccines included in the study were *Pneumococcal* vaccine, *Influenza* vaccine, *Typhoid* vaccine, *Varicella* vaccine, *Hepatitis A* vaccine and *MMR* vaccine. Following optional vaccines (with trade names) were included in the study: *Pneumococcal* vaccines (Prevenar-13, Synflorix); *Influenza* vaccine (Vaxigrip, Fluarix, Influvac); *Typhoid* vaccine (Typbar TCV, Typbar); *Varicella* vaccine (Variped, Varilrix); *Hepatitis A* (Havrix 720/1440, Avaxim 80, Biovac A); and *MMR* (Tresivac), as these were brands used in the study setup.

Government vaccine included for the study was *Measles* vaccine which also acted as a reference vaccine (control) for ensuring follow up of both genders equally post discharge after delivery. Government vaccines given during first 24 hours of birth like BCG, Hepatitis B and OPV were excluded from the study, since this would be dependent on institutional delivery load and will be biased by referral patients (who may not follow up for completing all their vaccines). DPT was excluded since it has been merged into pentavalent vaccines in the recent years, while rotavirus vaccine was excluded since it was introduced only in the recent years. Meningococcal Vaccine and Human Papilloma Virus Vaccine (though these are optional vaccines) are rarely used in this clinic and hence were excluded from the current study. Moreover, HPV Vaccine is usually given only to female children, and hence gender disparity cannot be assessed.

Data regarding the number of doses of the vaccine utilization of each month was collected using tally marks. Data was entered in Microsoft Excel Sheet 2016 version and analysed using Stata v 12.1 SE software licensed to one of the authors. Trend analysis plot was done using line graphs and difference in the number of doses by gender for each vaccine was analysed using independent sample t-test. Since the study did not include human subjects, and was record based, the researchers only informed the ethics committee regarding the same as per the mandate of the Institute Ethics Committee (IEC). No ethical approval was taken.

## RESULT

Data of vaccine dose utilization for 36 months (3 consecutive years) were collected from the clinic register. Total number of doses utilized, and that for male and female beneficiaries were noted for all vaccines considered in this study. Out of a total of 10,142 doses of vaccine utilized overall in this period, there were 5,313 (52.4 percent) vaccine doses given to male children and 4,829 (47.6 percent) doses to female. Of all optional vaccine doses given, *Pneumococcal* vaccine doses were the highest one utilized (19.6 percent) followed by *MMR* and *Typhoid* (Table 1). Considering all vaccines, on an average 281.72 ± 11.74 dose was utilized in a month with no statistically significant difference between male and female children (male 147.58 ± 6.74; female 134.13 ± 5.39; p = 0.123) (Table 2).

**Table 1:** Number of optional vaccine doses as per type and their proportion (n=10,142)

| Sl | Type of Vaccine | Number (%)<sup>^</sup> | Male (%) | Female (%) |
| --- | --- | --- | --- | --- |
| 1. | Pneumococcal | 1,988 (19.6) | 1,177 (59.2) | 811 (40.8) |
| 2. | Influenza | 599 (5.9) | 373 (62.3) | 226 (37.7) |
| 3. | Typhoid | 653 (6.4) | 420 (64.3) | 233 (35.7) |
| 4. | Varicella | 460 (4.5) | 210 (45.7) | 250 (54.3) |
| 5. | Hepatitis-A | 603 (5.9) | 360 (59.7) | 243 (40.3) |
| 6. | MMR | 735 (7.3) | 416 (56.6) | 319 (43.4) |
|  | All optional vaccines | 5038 (49.7) | 2956 (29.2) | 2082 (20.5) |
| 7. | Measles | 574 (5.7) | 302 (52.6) | 272 (47.4) |
|  | Total doses | 5612 (55.3) | 3258 (32.1) | 2354 (23.2) |

<sup>^</sup>Percentage is calculated from total number of doses for all vaccines i.e. 10,142 (excluding doses given at birth and BCG) out of which optional vaccines were 5,038(49.6%) over a period of 36 months

**Table 2:** Mean doses of each vaccine (total, and gender wise) received over last three years

| Sl | Vaccine | Mean ± SE<br>(Total doses) | Mean ± SE doses<br>(Male) | Mean ± SE doses<br>(Female) | P value (95% CI) |
| --- | --- | --- | --- | --- | --- |
| 1. | All Vaccines | 281.72 ± 11.74 | 147.58 ± 58 | 134.13 ± 5.39 | 0.123 (-3.77, 30.66) |
| 2. | All Optional Vaccines | 139.94 ± 8.12 | 82.11 ± 4.64 | 57.83 ± 3.68 | **0.000 (12.46, 36.09)** |
| 3. | Pneumococcal Vaccine | 55.22 ± 4.43 | 32.69 ± 2.63 | 22.52 ± 1.92 | **0.002 (3.65, 16.67)** |
| 4. | Influenza Vaccine | 16.63 ± 1.63 | 10.36 ± 1.01 | 6.27 ± 0.73 | **0.001 (1.58, 6.58)** |
| 5. | Typhoid Vaccine | 18.13 ± 1.37 | 11.66 ± 0.86 | 6.47 ± 0.58 | **0.000 (3.12, 7.26)** |
| 6. | Varicella Vaccine | 12.77 ± 1.48 | 5.83 ± 0.69 | 6.94 ± 0.86 | 0.320 (-3.32, 1.10) |
| 7. | Hepatitis-A Vaccine | 16.75 ± 0.92 | 10.00 ± 0.66 | 6.75 ± 0.36 | **0.000 (1.73, 4.76)** |
| 8. | MMR Vaccine | 20.41 ± 1.11 | 11.55 ± 0.69 | 8.86 ± 0.55 | **0.003 (0.92, 11.14)** |
| 9. | Measles Vaccine | 16.08 ± 1.00 | 8.38 ± 0.61 | 7.55 ± 0.45 | 0.280 (-0.69, 2.36) |

A total number of 5,038 (49.7 percent of all vaccines) optional vaccine doses were utilized during this period, with males receiving 2,956 doses (58.6 percent) and female children receiving 2,082 doses (41.4 percent). Mean number of total optional vaccines given per month was 139.94 ± 8.12, for males it was 82.1 ± 4.6 doses and for females it was 57.8 ± 3.7 doses. The mean number of total optional vaccine doses for males and females were found to be significantly different (p=0.0001, 95 percent CI: 12.46, 36.09) (Table 2), denoting that male children were receiving more number of optional vaccines in a month as compared to female children.

Among a total of 1,988 *Pneumococcal* vaccine doses utilized over this period, 1,177 doses were utilized for male and 811 doses for female children. The mean number of *Pneumococcal* vaccine dose used was 52.22 ± 4.44 per month, male receiving 32.69 ± 2.63, and female 22.52 ± 1.92 doses in a month. It was seen that male children received significantly more number of PCV doses per month than female children during this period (p=0.002, 95 percent CI: 3.65, 16.67) (Table 2). It was also found to be the most common optional vaccine opted for by parents.

*Influenza* and *typhoid* vaccine utilization had similar gender distribution. Out of a total of 599 *influenza* vaccine doses given during this period, 373 were for male and 226 for female children. The mean number doses were more for male as compared to female children (Male 16.63 ± 1.63; Female 10.36 ± 1.01; p = 0.001, 95 percent CI: 1.58, 6.58). Among a total of 653 *typhoid* vaccine doses, 420 doses were for male and 233 for female children. The mean number of total *typhoid* vaccine doses was 18.13 ± 1.37 per month, male children receiving 11.66 ± 0.86 and female 6.57 ± 0.58 doses. This was also found to be significantly more for male children (p = 0.000, 95 percent CI: 3.12, 7.26) (Table 2).

The case for *varicella* vaccine was interesting. A total of 460 doses had been utilized during this period out of which 210 number of doses accounted for male children vaccination in a month and 250 for female children. Here the mean number of total *varicella* vaccine doses was 12.77 ± 1.48 per month, 5.83 ± 0.69 for male children and 6.94 ± 0.86 for female. This was not found to be significantly different (p= 0.320, 95 percent CI: −3.32, 1.10) (Table 2).

There were 603 *hepatitis A* vaccine doses utilized during this period. Males had received 360 doses and females had received 243 doses in total. The mean number of doses received a month was 16.75 ± 0.92 and was significantly more for males as compared to females (Mean dose 10.0 ± SE 0.66 for male and 6.75 ± 0.36 for female, p = 0.000, 95 percent CI: 1.73, 4.76) (Table 2).

*Measles* vaccine, that is provided universally, was taken as the only government vaccine in this study. Males had received 302 and females 272 doses in total, out of 574 doses. The mean number of doses of *measles* vaccine for male and female children were not found to be statistically different (Male: 8.38 ± 0.61 doses/ month; female: 7.55 ± 0.45 doses/ month; p = 0.280, 95 percent CI: −0.69, 2.36) (Table 2). This vaccine acting as a control showed that there was no selection bias in the study, and in the long run there was an equal inflow of patients for both the gender among the study cohort receiving immunization. Comparison between *Measles* and *MMR* revealed interesting facts. The mean number of total doses for *MMR* vaccine was 20.41 ± 1.11 per month. Doses for male children were 11.55 ± 0.69 in a month while for female were 8.66 ± 0.55 doses a month, and this difference was found to be statistically significant (p = 0.003, 95 percent CI: 0.92, 11.14).

The trend of vaccination was found to be increasing in the immunization clinic of the hospital both in general and for optional vaccines (Fig 1 and 2). Individually all the optional vaccines showed a rising trend, while this was an exception for *measles* vaccine (Fig 3 to Fig 9). There was a downward trend for the total doses of *measles* vaccine received over the years (Fig 9), with a reverse trend (upward) for *MMR* vaccine (Fig 8). *Hepatitis A* vaccine and *MMR* showed a mild increasing trend while that for other vaccines was moderate to high, though this has not been quantified. The increasing trend was also evident in case of males and females for all the vaccines combined, as well as for PCV, *influenza, typhoid* and *hepatitis A*. Increasing trend was more for females than males for *Varicella* vaccine, and for *MMR* vaccine. There was even trend reversal with time in this case, with female children gradually taking up more vaccines as compared to males (Fig 6).

**Fig 1:**
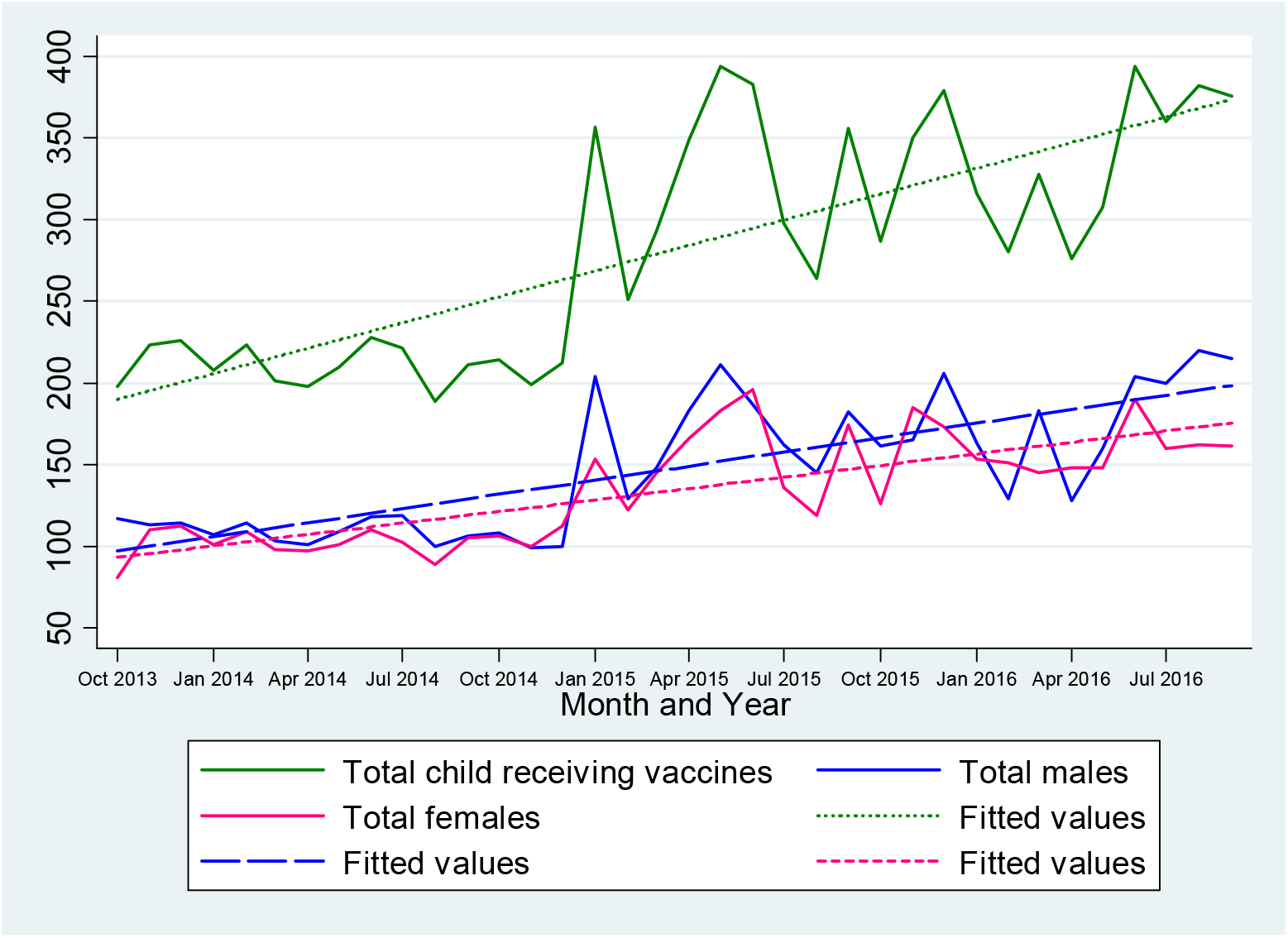
Trend of all vaccines utilized at the clinic from 2013 to 2016.

**Fig 2:**
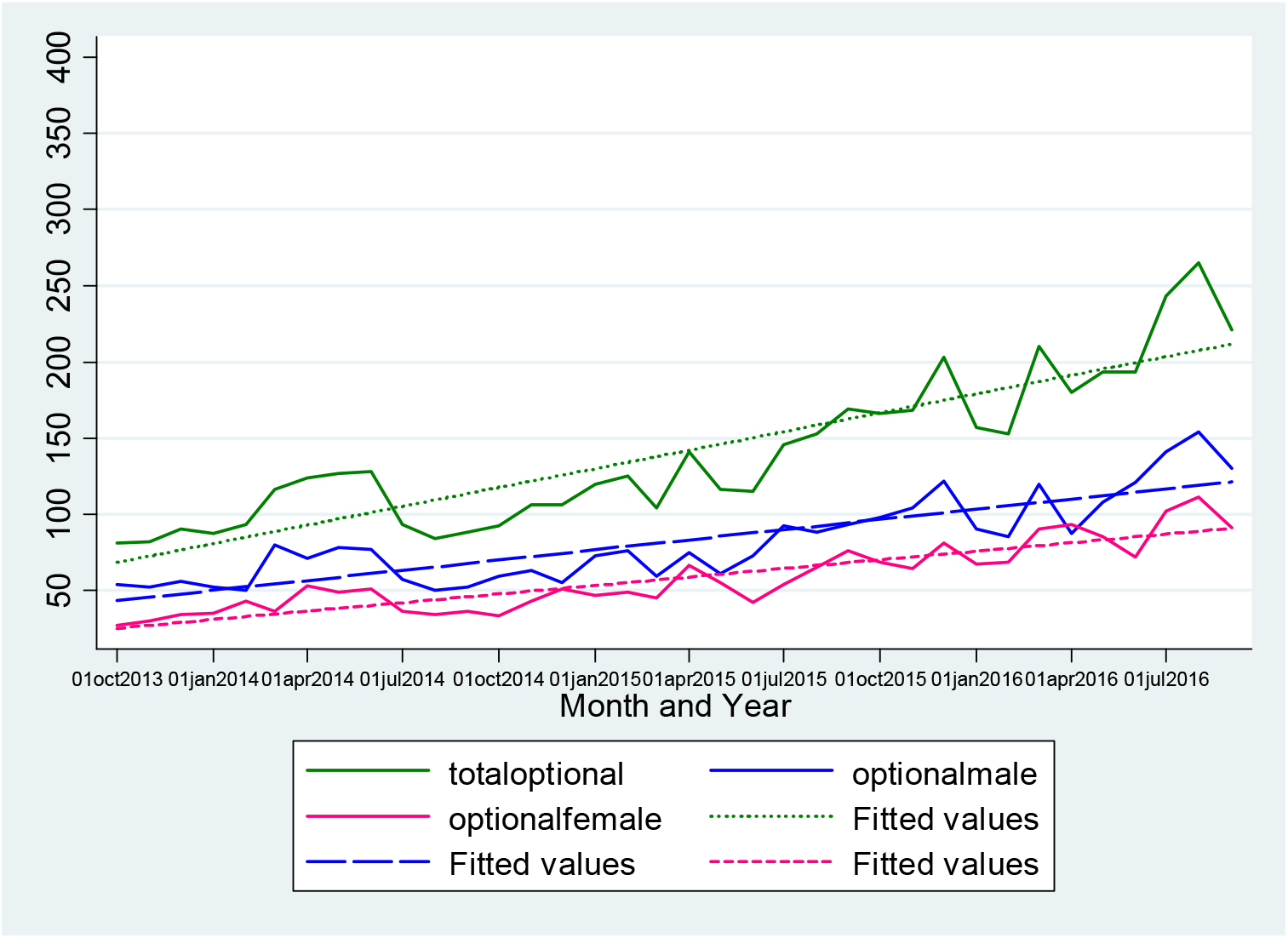
Trend of optional vaccines utilized at the clinic from 2013 to 2016.

**Fig 3:**
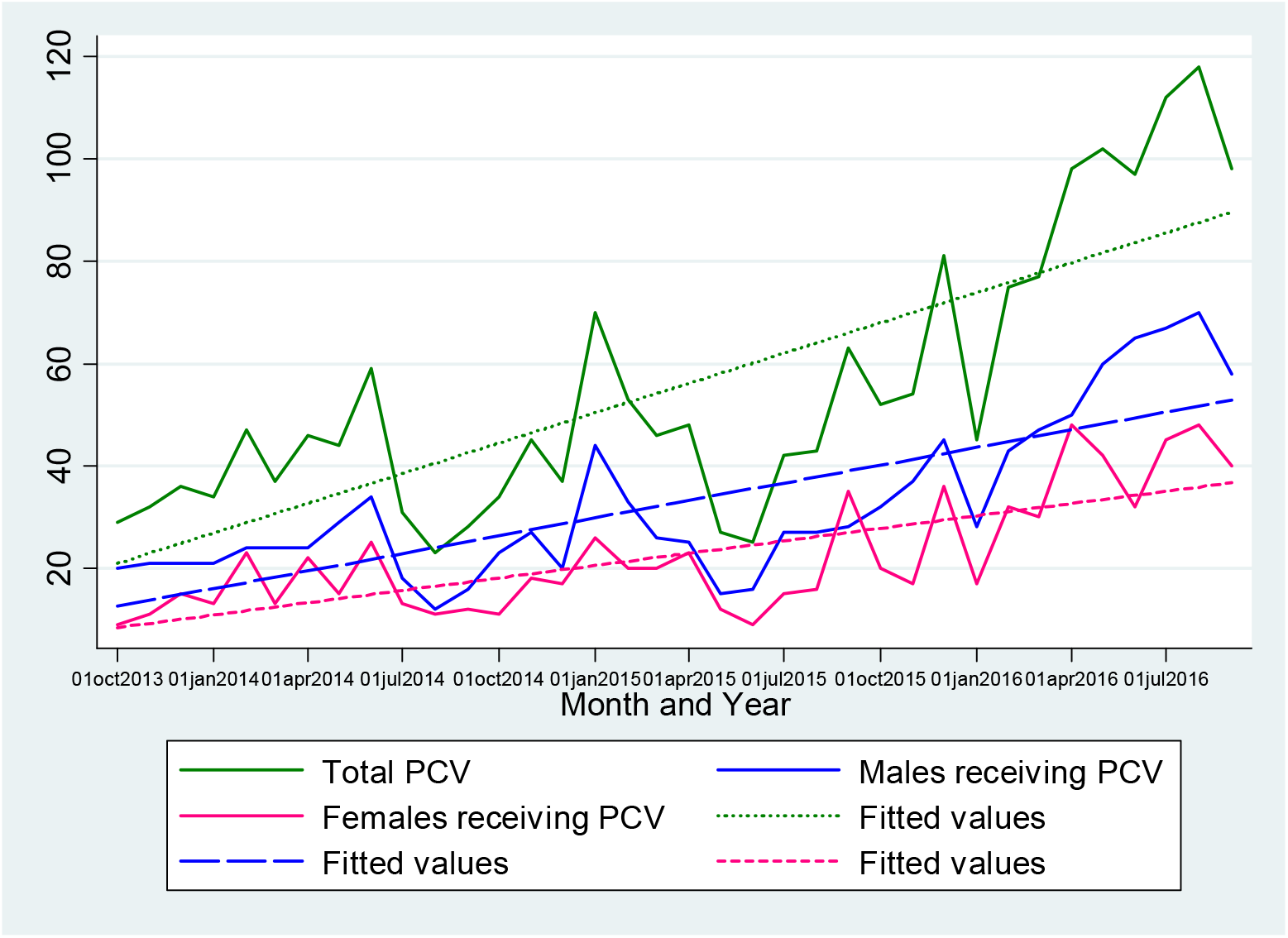
Trend of Pneumococcal vaccines utilized at the clinic from 2013 to 2016.

**Fig 4:**
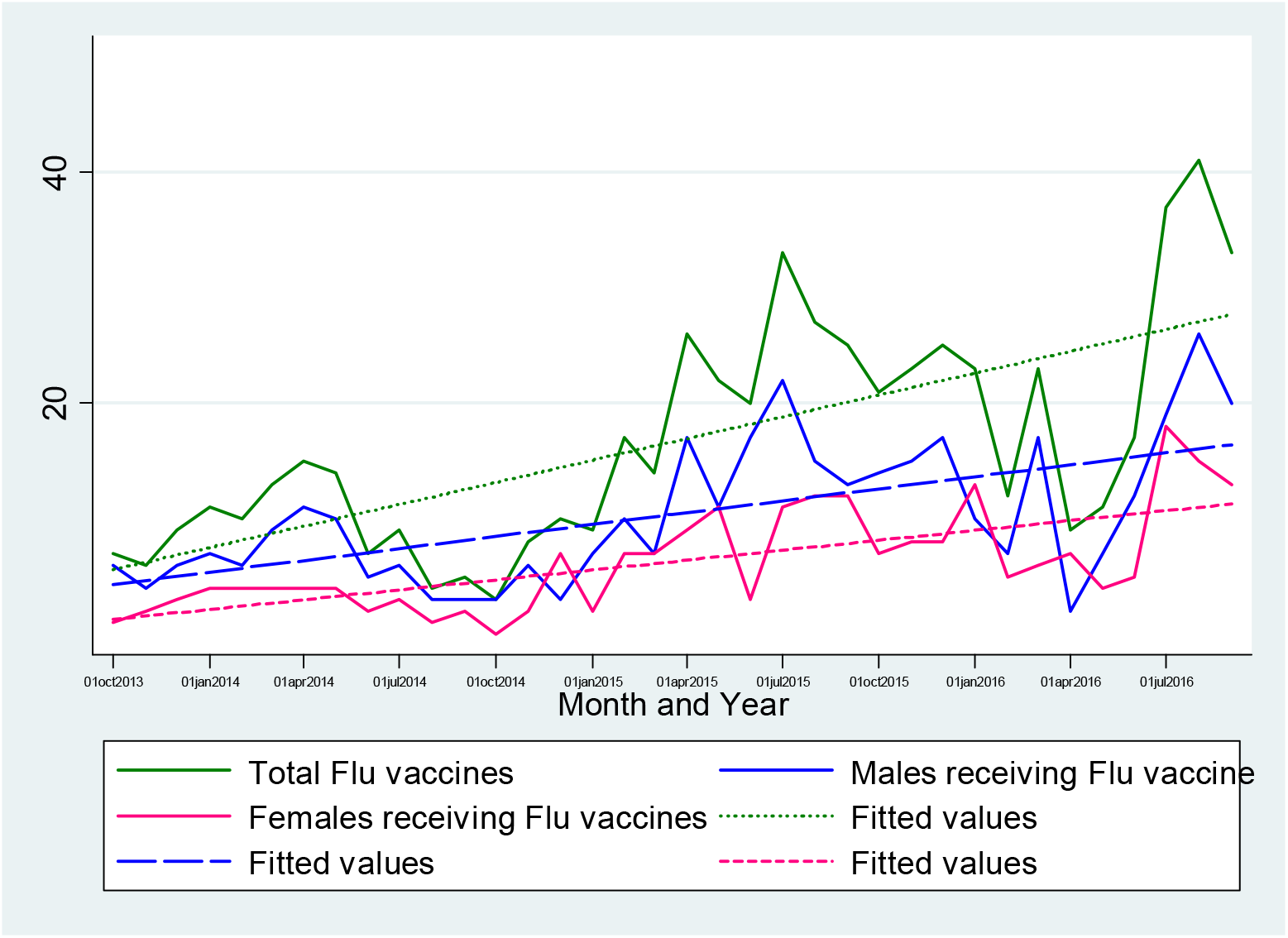
Trend of influenza vaccines utilized at the clinic from 2013 to 2016.

**Fig 5:**
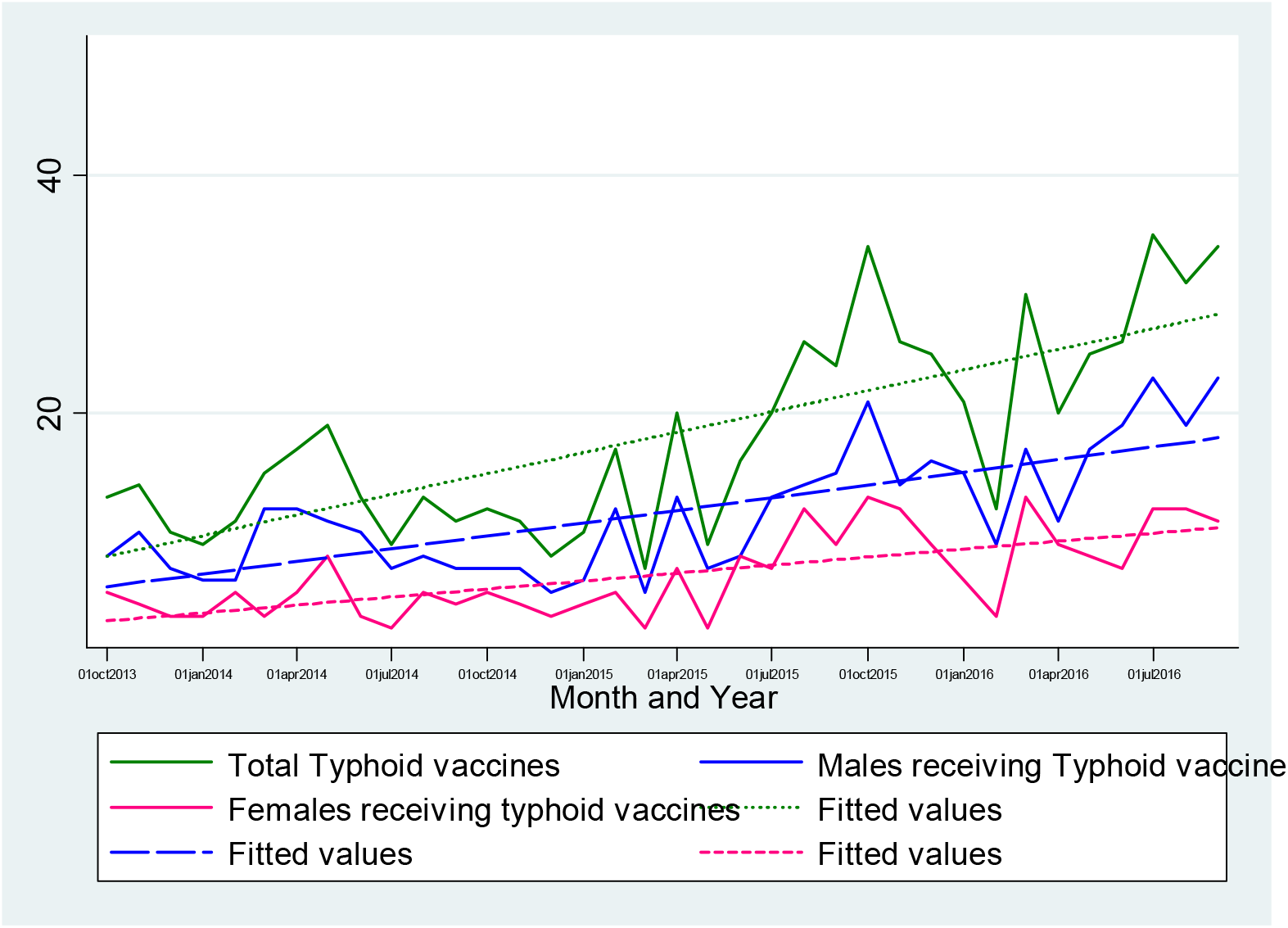
Trend of typhoid vaccines utilized at the clinic from 2013 to 2016.

**Fig 6:**
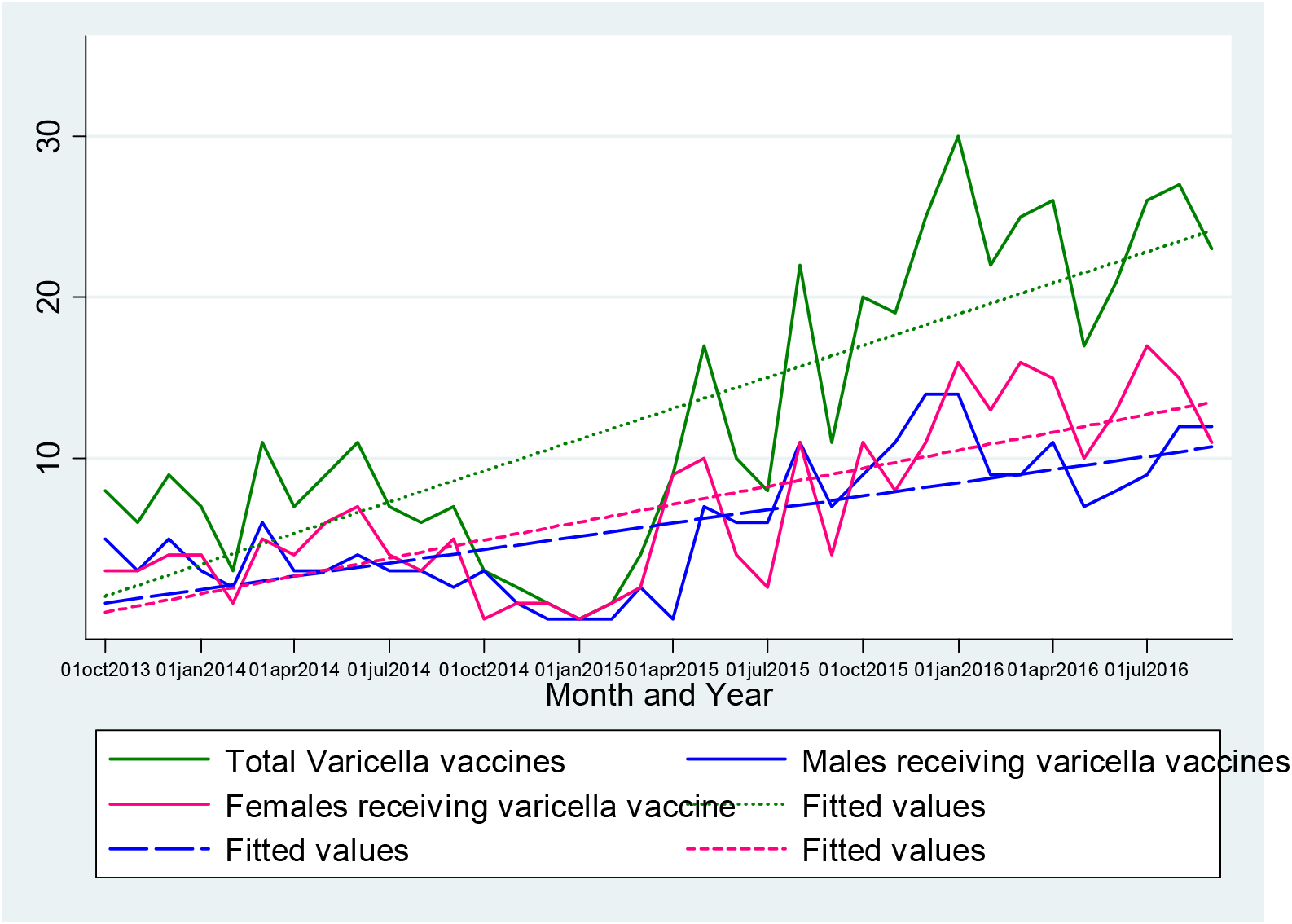
Trend of Varicella vaccines utilized at the clinic from 2013 to 2016.

**Fig 7:**
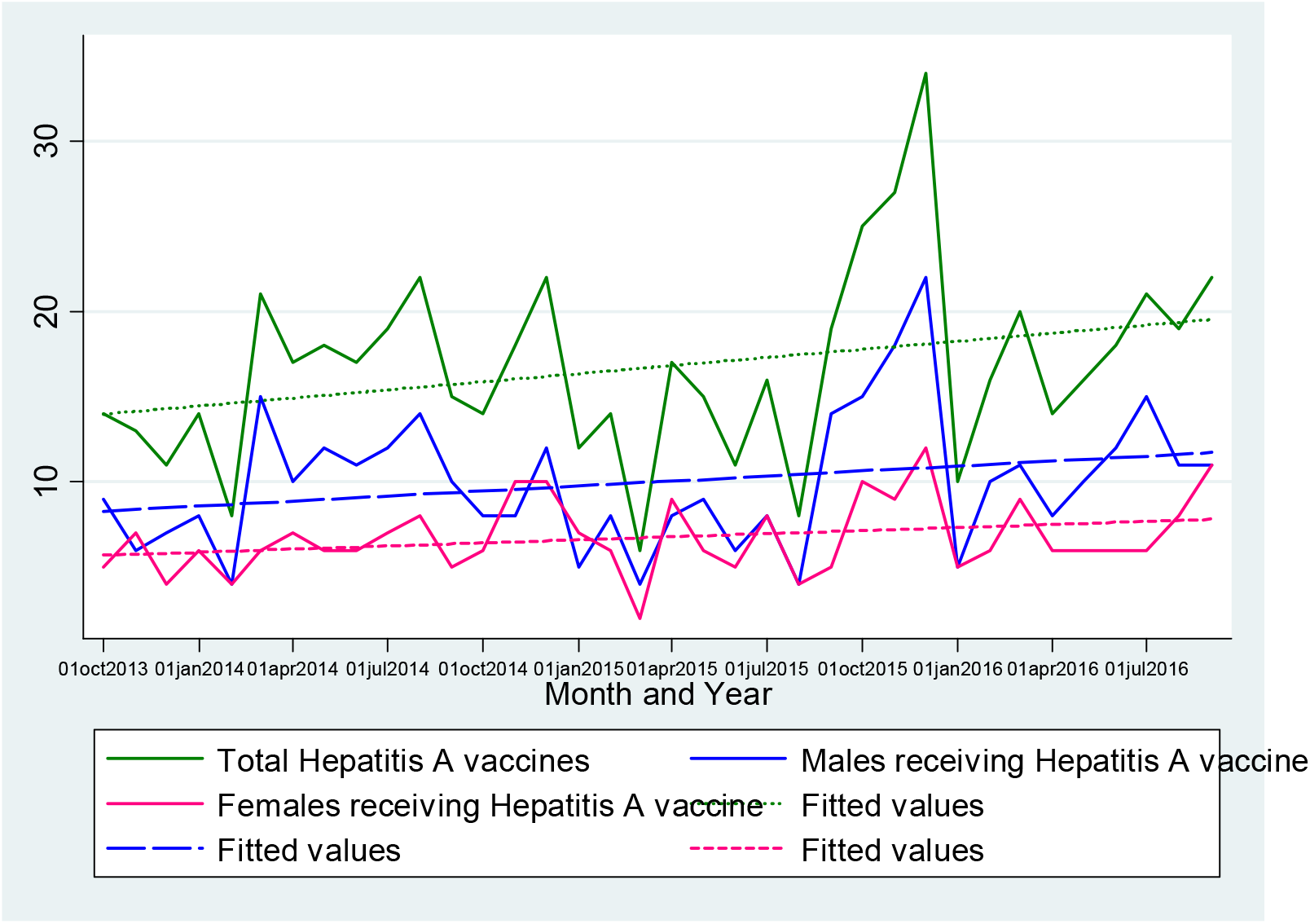
Trend of Hepatitis A vaccines utilized at the clinic from 2013 to 2016.

**Fig 8:**
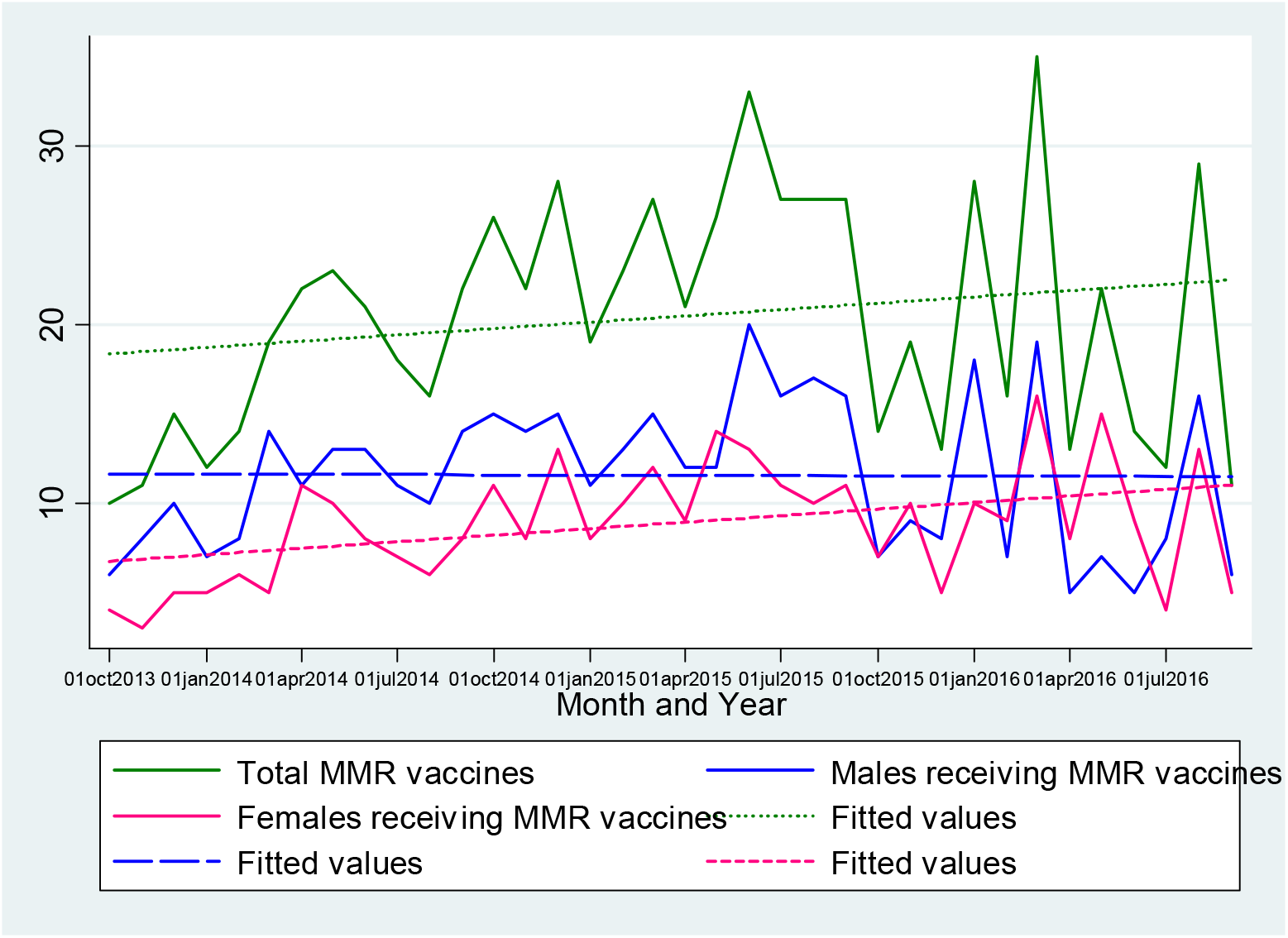
Total MMR vaccines utilized at the clinic from 2013 to 2016.

**Fig 9:**
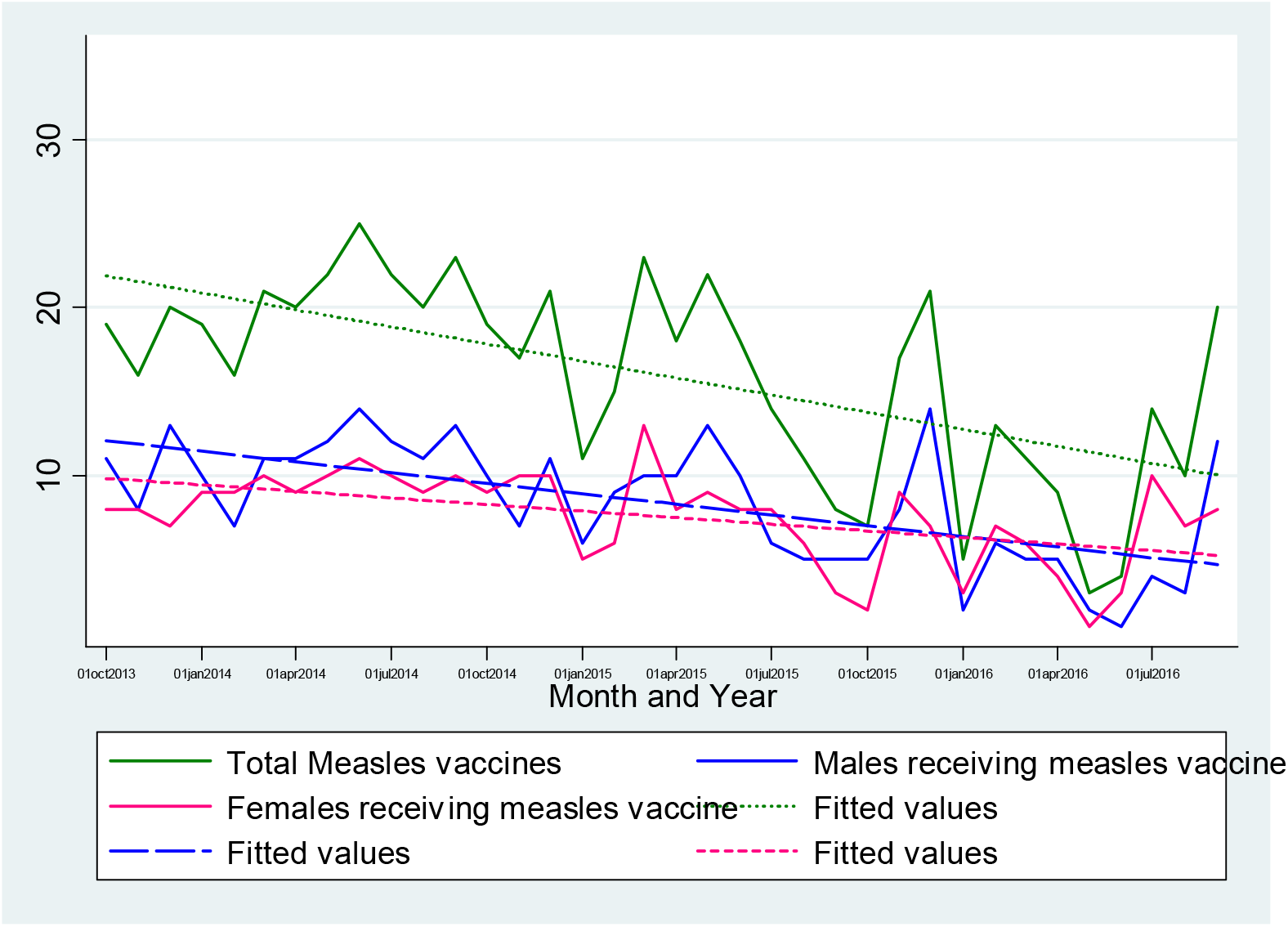
Total Measles vaccines utilized at the clinic from 2013 to 2016.

## DISCUSSION

Vaccines are estimated to prevent deaths in millions annually, and are of the greatest public health measures as on today. Starting from eradicating smallpox to the verge of eradicating polio and to a good extent measles, vaccination can be thought of as the most cost-effective public health intervention.(Delany, Rappuoli, and De Gregorio 2014)

For ensuring universal immunization coverage of all children below five years and also to eliminate maternal and neonatal tetanus from the country, the government of India is implementing Universal Immunization Program as fixed Routine Immunization(RI) sessions using platforms like Village Health and Nutrition Day(Panigrahi, Mohapatra, and Mishra 2015) or Routine Immunization Days. Vaccination is done using the National Immunization Schedule (NIS)(National Health Portal India 2018) for all children on every Wednesday at a village level. To increase urban immunization coverage (which many a times consists of floating population), National Urban Health Mission (a sub-mission of the National Health Mission, and aims to meet the health care needs of the urban poor, with special reference to the primary health care needs)(National Health Mission, Ministry of Health and Family Welfare, and Government of India 2020) also carries out similar activities. The sessions are usually conducted at the Anganwadi Centre. National Immunization Schedule is followed for the implementation of vaccination under this program. To increase immunization coverage and to reach out every individual, additional activities such as Mission Indradhanush (MI) and Intensified Mission Indradhanush (IMI) (to ensure full immunization coverage more than 90percent and reaching out the outreach areas) are also carried out.(NHP India 2018) Thus the government looks forward to avoid gender discrimination and uniformly vaccinate all children, mothers and adolescents. The effect of gender discrimination in case of vaccines under the national program thus seems to evade.

The Indian Academy of Paediatrics (IAP) has also recommended other additional vaccines such as *Pneumococcal, typhoid, hepatitis A, influenza, varicella, MMR, meningococcal* and *HPV* as optional vaccines as early as 2014.(Vashishtha et al. 2014) These optional vaccines are not financially supported by the government and they are only available to the individuals for purchase at private clinics or hospitals. Currently optional vaccines are being promoted and parents are being encouraged and counselled by the family practitioners and paediatricians to opt for these vaccines. Studies have shown that awareness among mothers regarding optional vaccines is around 66 percent. (Patnaik, Mishra, and Choudhury 2014). Thus optional vaccines, since not available free of cost, have the potential to show gender disparity, if present, in case of vaccination.

The optional vaccines used in this study were *Pneumococcal* vaccine, *Influenza* vaccine, *Typhoid* vaccine, *Varicella* vaccine, *Hepatitis A* vaccine and *MMR* vaccine. Following optional vaccines (with trade names) were included in the study: *Pneumococcal* vaccines (Prevenar-13, Synflorix); *Influenza* vaccine (Vaxigrip, Fluarix, Influvac); *Typhoid* vaccine (Typbar TCV, Typbar); *Varicella* vaccine (Variped, Varilrix); *Hepatitis A* (Havrix 720/1440, Avaxim 80, Biovac A); and *MMR* (Tresivac), as these were brands used in the study setup.

Pneumococcal vaccine used for the prevention of pneumococcal disease causes by *Streptococcus pneumoniae* bacteria, is commonly available as a PCV-10 (10-valent) or PCV- 13 (13-valent) vaccine under the trade name of Synflorix (one dose costing approximately 24 US$ in India) and Prevenar-13 (one dose costing approximately 51 US$ in India), respectively. PCV-13 contains serotypes for 1, 3, 4, 5, 6A, 6B, 7F, 9V, 14, 18C, 19A, 19F, and 23F pneumococcal strains. PCV10 contains all serotypes except 3, 6A, and 19A, although there is evidence for some cross-protection for 6A and 19A related disease. However, there is no potential advantage of using 13-valent over 10-valent as indicated in the study.(Temple et al. 2019) Thus, these two were used interchangeable for the current study. Another pneumococcal vaccine PPSV-23 is currently available which is a polysaccharide vaccine and recommended for geriatric population and people who smoke cigarettes or having any lung diseases like Chronic Obstructive Pulmonary Disease (COPD).(Centre for Disease Control and Prevention 2019b) PCV is currently given on pilot basis in certain districts of India in Universal Immunization Program, but is routine recommended as an optional vaccine by Indian Academy of Pediatrics.(Vashishtha et al. 2014; National Health Portal India 2018; National Health Mission 2018)

Influenza vaccine is a contagious disease which causes respiratory illness, and is very common in the west. The influenza season lasts from October to around mid-February (called as flu season). Persons at high risk of influenza include those above 65 years of age, with lung diseases like Asthma, diabetes, heart disease, etc., and even children in the young age. CDC recommends Flu vaccine routine to children once they cross six months of age to be given every year.(Centre for Disease Control and Prevention 2016b) However the recommendations differs in India, as per IAP. It is to be given for high risk category children, which in addition covers children having congenital, acquired or iatrogenic immunodeficiency, asplenia, renal or liver diseases, travellers, etc.(Vashishtha et al. 2014) In our setup, flu vaccine is a routine practice given to all children above six months of age as per CDC guidelines, and then every year. Flu vaccine is available as Vaxigrip (Sanofi India; costing approximately 12 US$), Fluarix (Glaxo Smith Kline Limited; costing approximately 10 to 14 US$) and Influvac (Abbott Pharma; costing approximately 13 US$ in India). Flu vaccine covers usually the commonly circulating strains of both type A and type B influenza. Below 3 years of age, 0.25 ml dose is given and above that 0.5 ml as per posology and indication of the brand.(MIMS 2020; Grohskopf et al. 2016; “Vaccines - Sanofi India” 2019) Typhoid vaccine helps in preventing typhoid fever, which may be life threatening. Earlier a live form (above 6 years and given every 5 years) and an inactivated form (above two years and to be given every two years) was available. Typhoid vaccine is routinely not recommended in US (only to be given for travellers, lab handlers or close contacts of typhoid cases/ carriers) but in India recommended by Indian Academy of Pediatrics in the age group of 9 to 12 months. Usually the conjugate vaccine is in use and available in Indian market.(Vashishtha et al. 2014; US Department of Health and Human Services and CDC 2019) It is available as Typbar TCV (Bharat Biotech; costing approximately 24 US$ in India), and it a vaccine containing Salmonella typhi Ty2 polysaccharide. Tetanus Toxoid is used as a conjugate and helps in a lasting immunity even without booster doses (single dose eliciting seroconversion in 98.05 percent).(Bharat Biotech 2019) Typhoid conjugate vaccine is given by intramuscular route after two years of age (now can be given in less than 2 years also) with a dose of 0.5 ml.

Varicella Zoster vaccine (VZV) is a vaccine for prevention of the very contagious disease called Chicken-pox which causes blisters with itching, fever and tiredness. Vaccine available for the same have been reviewed by World Health Organization for its effectiveness and concluded that even with a single dose given to protect children of 9 months to 12 years against all grades of severity of varicella disease, an approximate mean vaccine efficacy was 80 percent against all grades of disease severity, irrespective of vaccine type. Two doses of the vaccine whereas was said to be effective in about 90 percent of cases for preventing chicken pox. VZV is available in India as Varilrix (Glaxo Smith Kline Limited; costing approximately 16 US$ in India) and Variped (Merck Sharp and Dohme Corporation India; costing approximately 19 US$ in India), most commonly. They are given 0.5 ml by subcutaneous route. (Centre for Disease Control and Prevention 2016a; Strategic Advisory Group of Experts on Immunization 2014)

Hepatitis A is a form of hepatitis caused by the hepatitis A virus (HAV), which can affect anyone. Hepatitis A vaccine is effective for the long-term prevention of HAV infection and can be given to persons one year of age and older. Hepatitis is transmitted through feco-oral route and India has seen outbreaks related mostly to Hepatitis A or Hepatitis E, at various points of time.(Paul et al. 2015; Kumar et al. 2015; Rakesh et al. 2014) This may be also the reason why IAP recommends vaccination of children with Hepatitis A vaccine from one year of age.(Vashishtha et al. 2014) Hepatitis A vaccine is available in India as Havrix junior 720 (Glaxo Smith Kline Biologicals; approximate cost is 15 US$) or Avaxim 80 (Sanofi India Limited; approximate cost is 18 US$) and is given in children as two doses of 0.5 ml each 6 months apart by intramuscular route. Alternate to this inactivated vaccine is a single dose of live vaccine available as Biovac A (Wochardt Limited; approximate cost of one dose is 17 US$) given subcutaneously as 0.5 ml. All have a good seroconversion rate of more than 95 percent if given as per schedule. (Bhave et al. 2015; Abarca et al. 2008)

Measles, Mumps and Rubella are viral contagious diseases. Measles is caused by Measles virus, and causes fever with rash along with cough, runny nose, and red, watery eyes. Diarrhoea is a usual complications along with pneumonia, and death. Mumps is caused by mumps virus and leads to inflammation of salivary glands (mainly parotid). It is accompanied by fever, headache, muscle aches, tiredness and loss of appetite. Encephalitis/meningitis may be a complication. Rubella (also known as German Measles) causes fever, sore throat, rash, headache, and red, itchy eyes. If infected during pregnancy it may lead to a miscarriage or baby born with serious birth defects. Hence children 12 through 15 months of age are to be vaccinated with 0.5 ml subcutaneous dose, with a booster dose at 5 to 6 years of age. (Centre for Disease Control and Prevention 2019c, 2019a) MMR is a live attenuated vaccine, and is very economic to be afforded by individuals in the developing countries. In India it is marketed as Tresivac (Serum Institute India Pvt. Ltd.; cost for one dose approximately 1 US$).(Serum Institute of India Pvt. Ltd. 2019)

*Pneumococcal* vaccine was the most common optional vaccine (highest number of doses) used by parents as per this study. The reason may be because it was given on multiple occasions as 3+1 or 2+1 schedule for an under five child. From this study, we found that there was an increasing trend in all vaccines. Optional vaccines taken overall and individually *(Pneumococcal* vaccine, *Influenza* vaccine, *Typhoid* vaccine, *Varicella* vaccine, *Hepatitis A* vaccine and *MMR* vaccine) showed a strong increasing trend, with mild increasing trend seen only for *Hepatitis A* vaccine. *Measles* vaccine showed a decreasing trend. This increasing trend could be due to increasing awareness and acceptability of parents for optional vaccines. There was gender disparity (preference for male children) observed for all the optional vaccines except *varicella* vaccine among optional vaccines and *measles* vaccine among vaccines supplied under Universal immunization program (UIP).

A study done by Padda et al. in the community setting in other parts of India has also shown that the coverage of optional vaccines was significantly more for males as compared to females (53.9 percent for male vs 46.1 percent for female).(Padda et al. 2012) A study done for the analysis of factors responsible for uptake of rotavirus vaccine in a paediatric clinic also showed association of gender of the baby with opting for optional vaccines. Other factors associated were mother's age and occupation, the mode of payment, the number of previous visits and counselling sessions and the pre-counselling awareness or knowledge were significantly associated with uptake of vaccine.(Kutty et al. 2010)

For every optional vaccine that were meant to purchase by the parent, preference was given to male children over female excepting for *varicella* (chicken pox) vaccine. Vaccines that were provided free of cost by the government under immunization program had no gender difference in vaccine utilization (evident from comparison of gender disparity for *measles* vaccine as control). For *MMR* vaccine gender disparity was present in favour of male children, even though the cost of this vaccine was reasonably low in comparison, and even when it was of public health importance in countries like India when given to female children.

*Varicella* vaccine was found to be an exception where there was no difference in vaccine dose utilization between male and female children. This may be probably due to social stigma for delay in marriage of their girl child due to scar marks on face and elsewhere arising out of chicken pox, but has not been ascertained in this study.

Finally, there was no single optional vaccine in our study that was provided more to female as compared to male children. Further qualitative research might be needed to find out the reasons behind this disparity that are amenable to corrections. By doing qualitative research probably we will get to know many themes(Angelillo et al. 1999; Patnaik, Mishra, and Choudhury 2014) like lack of knowledge of parents about the importance of vaccination for female children; importance of various socio-demographic factors leading to gender disparity; education of the decision maker in the family for financial aspects (usually father); target for health education (father/ mother/ both); or approaches in communication for health education.

The study could have been biased by the bias in counselling technique of the physicians in the immunization and health guidance clinic, but this was avoided by certain principles. One, there was uniform displays of information, education and communication materials. Second, there was a strict protocol from entry of the beneficiary to the exit of the beneficiary and every physician joining the institute was trained and made versed with the protocol before getting posted. Third, there was an independent counselling corner for resolving every small query of the client. The next bias could have been marked differences in pricing of the optional vaccines. But as mentioned earlier, the pricing difference was very minimal except for the brands of pneumococcal vaccine (10-valent costing quite less than 13-valent PCV). However, the beneficiaries were given choices to select even PVC-10 over PCV-13 in case of having financial constraints. Analysis was done merging both as PCV and not independently, thus avoiding bias during analysis.

Gender inequality has been a tradition in Indian culture. Female feticide is trend reported in Asian countries, and prevalence to a large extent in many regions of India, especially in northern India where the child sex ratio drops to less than 850 over the country average of 914 as per Census 2011.(Ahmad and Darussalam 2010; Office of the Registrar General of India 2011a) A higher proportion of under five female children with birth order two or more, and belonging to anganwadis of urban slums of Maharashtra (India), were found to be malnourished compared to the male children.(Patel et al. 2013) The national difference in the male to female literacy rate still stands at 16.68 points (male 82.14 percent, female 65.46 percent), though has been reduced than the previous Census in 2001. The difference is marked in cases of Empowered Action Group States (EAG), which contribute to high burden of disease in the country.(Office of the Registrar General of India 2011b) Moreover there has been an Indian Psychology attached at every stage of life in India, existing not only in the rural and uneducated population of the country, but also in other sections also. Adolescent girls in the country are imparted less knowledge on sex education, making sometimes menstrual hygiene also difficult to maintain. Early school drop out followed by early marriage, and early child bearing is a custom very common in rural India. Ignorance of legal rights by a girl also leads to issues like exposure to domestic violence, economic dependence, low self-esteem, no rights on property, denial of decision making rights, child immoral trafficking and exposure to sexually transmitted diseases, etc.(Rao, Vidya, and Sriramya 2015)

Continuous efforts have been taken up by the Indian government to demolish this discrimination for female supported by global developmental partners. Pre-conceptional and pre-natal diagnostic (PC-PNDT) act 1994 (amended in 2011) prohibits on sex determination at the time of delivery and even during conception (during artificial insemination).(Bhaktwani 2012) Inclusion of education on sexual and reproductive health as a part of Reproductive Maternal Neonatal Child Health Plus Adolescent since 2014 (RMNCHA Strategy) where Rashtriya Kishor Swasthya Karyakram (RKSK) is rolled out for adolescents.(National Health Mission 2020) Similarly, universalization of immunization ensures that a girl child even gets vaccinated for the common vaccine preventable diseases. Laws have been imposed to restrict teenage marriage and pregnancy. Girl education has also been prioritized through initiatives like Beti Bachao Beti Padhao Scheme by the Ministry of Women and Child Development. (Ministry of WCD Government of India 2017) Apart from this, government has taken many steps to remove the gender bias towards a girl child, like reservation of seats in technical education, legislative assemblies of parliament, government jobs, etc.(Rao, Vidya, and Sriramya 2015)

However, in spite of all the measures taken up by the government, this study revealed an interesting untouched area regarding differences in use of optional vaccines, which points towards the still existing gender differences. This can be considered as a proxy indicator in the assessment of gender differences alongside with other indicators taken in Census or other surveys. Gender Inequality Index (GII) is commonly used for measuring gender differences, and hovers around three key components like health, empowerment and labour market, based on which female gender index and male gender index are calculated. (United Nation Development Programme 2019) But considering a small geographical area or a state/ county, an indicator like this – use of optional vaccines among the population – can show the gender differences still existing in the field of health, more related to attitude of the population.

Organizations like the Global Alliance for Vaccine Initiative (GAVI)(WHO 2016), also exert substantial pressure on developing countries to incorporate these optional vaccines into their immunization schedule. Recently there are pilot projects ongoing for introduction of Pneumococcal conjugate vaccine (PCV), Measles Mumps and Rubella (MMR) trivalent vaccine and even Japanese-B Encephalitis (JE) vaccine.(Paul and Sahoo 2015; Mishra and Mishra 2008) Injectable Polio Vaccine (IPV) has also been incorporated in the schedule and given at cold chain points as fractional dose of IPV given intradermal at 6 and 14 weeks, though initially planned for a single dose at 14 weeks.(World Health Organization 2019; Indian Academy of Pediatrics (IAP) Advisory Committee on Vaccines and Immunization Practices (ACVIP) et al. 2016) But at every point of time, there will be additional vaccines made available in the market, which are not in the immunization program of the country, which can act as proxy indicator to point at the health related gender discrimination in the community.

## CONCLUSION

From this study, we concluded that there was an observable increasing trend in vaccination at this tertiary care facility. Gender disparity was probably present in all the optional vaccines except for *varicella* vaccine. A well-designed cohort study may throw more light on this issue. More robust counselling may be needed whenever there is a girl child which may contribute to reduced gender disparity.

## LIMITATION

1. This is a record based study, hence there may not be strong level of evidence.
2. Most of the sociodemographic characteristics of parents were not captured, which were beyond the capacity of the researchers.
3. Doses of vaccine have been considered and not children as cohort. Thus, different children might have received the doses.
4. The findings are limited to a tertiary care facility, and may need interpretation with care when considered for generalizing.
5. Counselling technique of the clinicians have not been taken into consideration, which might influence the uptake of optional vaccines thus leading to gender disparity.

## Data Availability

Data will be made available on request to the corresponding author

